# Effects of XFG-adapted mRNA vaccination on SARS-CoV-2 variant neutralisation

**DOI:** 10.64898/2026.10.04.26364675

**Authors:** Christine Happle, Markus Hoffmann, Metodi V. Stankov, Nianzhen Chen, Inga Nehlmeier, Luise Graichen, Anna-Sophie Moldenhauer, Torsten Witte, Stefan Pöhlmann, Alexandra Dopfer-Jablonka, Georg M.N. Behrens

## Abstract

SARS-CoV-2 antigenic evolution continues to challenge strain-matched vaccination. For the 2026–27 season, the European Medicines Agency (EMA) identified XFG as the preferred antigen within the JN.1 family and Comirnaty XFG was authorised in the European Union in July, 2026. We studied 22 participants who received 30 μg of mRNA encoding XFG (Comirnaty XFG, BioNTech–Pfizer, Mainz, Germany) in September, 2026 and had paired serum available before vaccination and median 14 days afterwards. Median Wuhan-Hu-1 S1-specific IgG increased from 1600 BAU/mL (IQR 1118–2926) to 2914 BAU/mL (2117–4468), a 1·8-fold increase. Median RBD-binding ED50 increased from 116 (51·4–212) to 305·6 (170·8–434·6) for LP.8.1 and from 354·3 (129·2–558·2) to 832·9 (429–1175) for NB.1.8.1, corresponding to 2·6-fold and 2·4-fold increases, respectively (all p<0·0001). XFG-adapted vaccination increased neutralising activity against each tested variant. The GMT against XFG increased from 180 before vaccination to 894 afterwards (5·0-fold), and that against RV.1.3 increased from 121 to 599 (4·9-fold). GMTs increased from 117 to 386 for PQ.16.1.1 (3·3-fold), from 101 to 316 for XFG.1.1 (3·1-fold), from 629 to 1772 for LP.8.1 (2·8-fold), from 221 to 611 for NB.1.8.1 (2·8-fold), and from 228 to 642 for PJ.2.1 (2·8-fold). Neutralisation of BA.3.2.2 increased more modestly, from GMT 78 to 126 (1·6-fold; p=0·0009). Pre-versus post-vaccination neutralisation titres differed significantly for all variants (p<0·001). Despite the broad increase, post-vaccination neutralisation remained heterogeneous. Relative to LP.8.1, GMTs were 2·0-fold lower for XFG, 2·8-fold lower for PJ.2.1, 2·9-fold lower for NB.1.8.1, 3·0-fold lower for RV.1.3, 4·6-fold lower for PQ.16.1.1, 5·6-fold lower for XFG.1.1, and 14·1-fold lower for BA.3.2.2. These findings provide early human immunogenicity data after XFG-adapted mRNA vaccination. The largest relative increases were observed for XFG and RV.1.3, whereas LP.8.1 remained the most strongly neutralised variant after Comirnaty XFG vaccination and BA.3.2.2 the least. This pattern suggests broad boosting of cross-reactive humoral immunity without elimination of between-variant differences. The small, extensively pre-immunised cohort, short follow-up, absence of a contemporaneous vaccine-comparator group, and use of a pseudovirus assay limit inference. Our data characterise early antibody binding and neutralisation but do not establish durability or clinical effectiveness.

## Dear Editor

SARS-CoV-2 antigenic evolution continues to challenge strain-matched vaccination. For the 2026–27 season, the European Medicines Agency (EMA) identified XFG as the preferred antigen within the JN.1 family and Comirnaty XFG was authorised in the European Union in July, 2026 (1,2). EMA emphasised the need for post-authorisation immunogenicity and effectiveness data. We therefore characterised early humoral responses after XFG-adapted mRNA vaccination against XFG and a panel of contemporary SARS-CoV-2 lineages.

We studied 22 participants in the prospective COVID-19 Contact (CoCo) study who received 30 μg of mRNA encoding XFG (Comirnaty XFG, BioNTech–Pfizer, Mainz, Germany) in September, 2026 and had paired serum available before vaccination and 11–16 days afterwards (median 14 days; appendix p2). Their median age was 49 years (range 35–77), 11 (50%) were male, all had previously received COVID-19 vaccines, and 19 (86%) reported at least one previous SARS-CoV-2 infection. We quantified spike-binding IgG and neutralisation of vesicular stomatitis virus-based pseudoviruses bearing spike proteins from LP.8.1, BA.3.2.2, NB.1.8.1, PQ.16.1.1, XFG, XFG.1.1, RV.1.3, and PJ.2.1 (appendix p7).

Binding-antibody responses increased after vaccination (Figure A). Median Wuhan-Hu-1 S1-specific IgG increased from 1600 BAU/mL (IQR 1118–2926) to 2914 BAU/mL (2117–4468), a 1·8-fold increase. Median RBD-binding ED50 increased from 116 (51·4–212) to 305·6 (170·8–434·6) for LP.8.1 and from 354·3 (129·2–558·2) to 832·9 (429–1175) for NB.1.8.1, corresponding to 2·6-fold and 2·4-fold increases, respectively (all p<0·0001). Binding to the corresponding S1+S2 proteins increased 1·9-fold and 2·3-fold (appendix p8).

**Figure:**
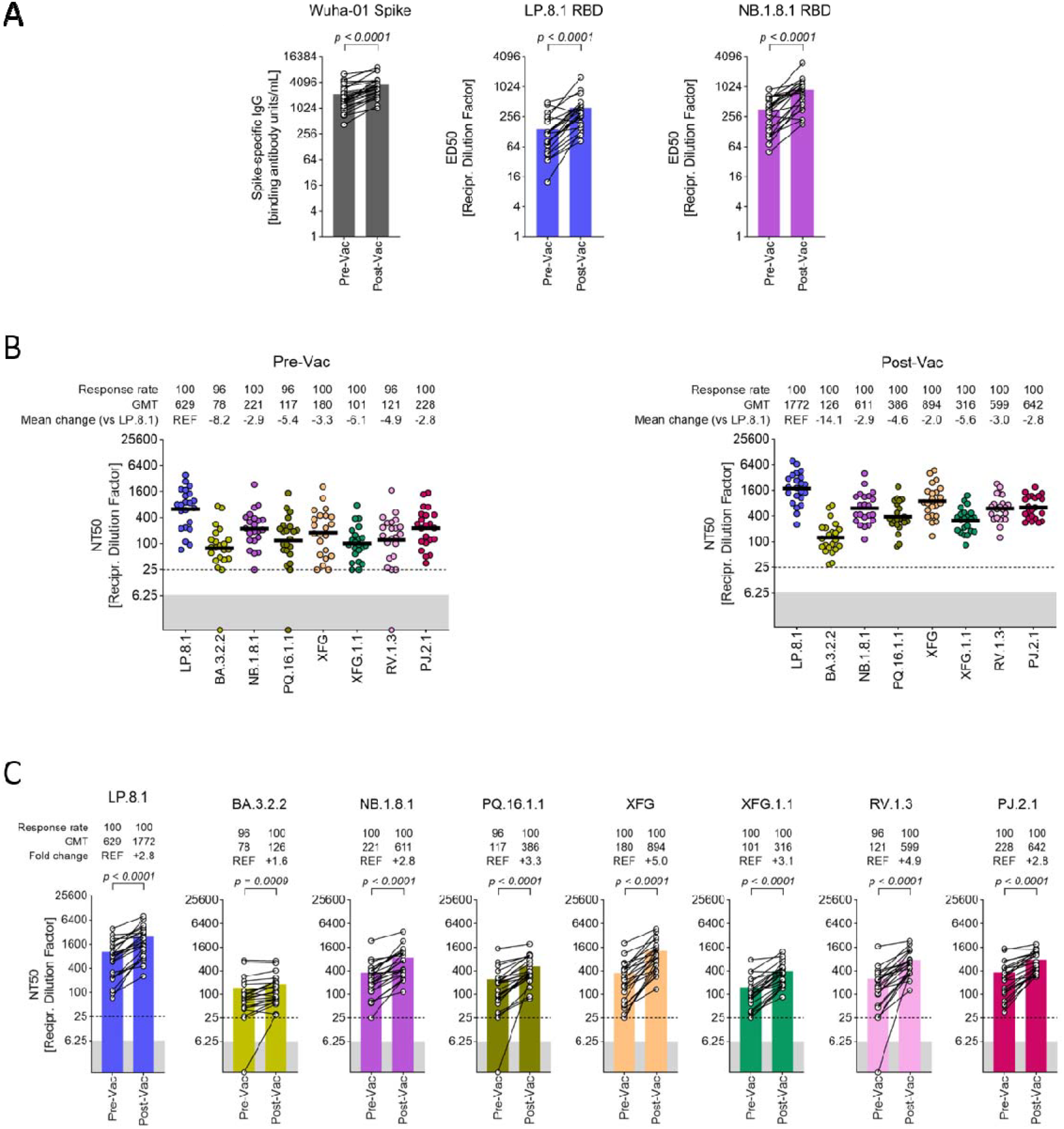
Humoral immune responses after XFG-adapted mRNA vaccination. (A) Wuhan-Hu-1 S1-specific IgG and RBD-binding ED50 titres against LP.8.1 and NB.1.8.1 before and after vaccination (n=22). Paired comparisons used two-sided Wilcoxon matched-pairs signed-rank tests. (B) Neutralisation of vesicular stomatitis virus-based pseudovirus particles bearing the indicated S proteins by donor-matched serum (n=22). Data are grouped to compare differences in SARS-CoV-2 lineage-specific neutralisation before and after omicron XFG vaccination. Information on GMT (also indicated by horizontal lines), response rates, and mean fold changes in neutralisation compared with LP.8.1 pseudovirus particles are indicated below the graphs. (C) Data in (B) were regrouped to compare differences in SARS-CoV-2 lineage-specific neutralisation before and after vaccination. Information on response rates, GMT (also indicated by coloured bars), and mean fold changes in neutralisation from before to after vaccination are indicated below the graphs. Data represent GMTs from a single experiment, performed with four technical replicates. The lowest serum dilution tested (dashed lines) and the threshold (lower limit of detection; grey shaded areas) are indicated. Of note, for graphical reasons, serum samples yielding an NT50 value less than 6·25 (limit of detection) were manually set at the bottom of the axis. Individual neutralisation data are available in the appendix (pp 9–10). P values indicates statistical significance by Wilcoxon–Mann–Whitney test. Circles indicate individual participants. GMT=geometric mean titre. NT50=50% neutralising titre.

XFG-adapted vaccination increased neutralising activity against each tested variant (Figure B, C). The GMT against XFG increased from 180 before vaccination to 894 afterwards (5·0-fold), and that against RV.1.3 increased from 121 to 599 (4·9-fold). GMTs increased from 117 to 386 for PQ.16.1.1 (3·3-fold), from 101 to 316 for XFG.1.1 (3·1-fold), from 629 to 1772 for LP.8.1 (2·8-fold), from 221 to 611 for NB.1.8.1 (2·8-fold), and from 228 to 642 for PJ.2.1 (2·8-fold). Neutralisation of BA.3.2.2 increased more modestly, from GMT 78 to 126 (1·6-fold; p=0·0009). Pre-versus post-vaccination neutralisation titres differed significantly for all variants (p<0·001).

Despite the broad increase, post-vaccination neutralisation remained heterogeneous. Relative to LP.8.1, GMTs were 2·0-fold lower for XFG, 2·8-fold lower for PJ.2.1, 2·9-fold lower for NB.1.8.1, 3·0-fold lower for RV.1.3, 4·6-fold lower for PQ.16.1.1, 5·6-fold lower for XFG.1.1, and 14·1-fold lower for BA.3.2.2. The low BA.3.2.2 neutralisation is consistent with previous reports of marked antigenic separation of BA.3.2 from contemporary JN.1-derived variants (3,4). Conversely, a recent preprint reported that PJ.2.1 combines markedly increased ACE2-binding capability with antigenic similarity to the JN.1 family and no strong humoral immune evasion (5), consistent with the comparatively preserved PJ.2.1 neutralisation in our assay.

These findings provide early human immunogenicity data after XFG-adapted mRNA vaccination. The largest relative increases were observed for XFG and RV.1.3, whereas LP.8.1 remained the most strongly neutralised variant after Comirnaty XFG vaccination and BA.3.2.2 the least. This pattern suggests broad boosting of cross-reactive humoral immunity without elimination of between-variant differences. The small, extensively pre-immunised cohort, short follow-up, absence of a contemporaneous vaccine-comparator group, and use of a pseudovirus assay limit inference. Our data characterise early antibody binding and neutralisation but do not establish durability or clinical effectiveness.

During the preparation of this work the authors used ChatGPT (OpenAI) and Claude (Anthropic) in order to assist with grammar and style corrections. After using this service, the authors reviewed and edited the content as needed and take full responsibility for the content of the publication.

## Data Availability

All requests for raw and analysed data that underly the results reported in this article will be reviewed within four weeks by the CoCo Study Team, Hannover Medical School to determine whether the request is subject to confidentiality and data protection obligations. Data that can be shared will be released via a material transfer agreement.

