## Appendix for "Effects of XFG-adapted mRNA vaccination on SARS-CoV-2 variant neutralisation"

**Content** **Page**

Table S1. Demographics and infection and vaccination history 2

Methods 2

Limitations of the study 4

Acknowledgements and declarations 5

Supplementary references 6

Supplementary figures 7

**Table S1. Demographics and infection and vaccination history**

| Variable | Vaccinees |
|---|---|
| Number of vaccinees | 22 |
| Age, Median (IQR), range | 49 (15), 35-77 |
| Sex, male (%) | 11 (50) |
| Prior COVID-19 vaccination (%) | 22 (100) |
| Number of COVID-19 vaccinations: Median (IQR), range | 7 (2), 4-11 |
| Months since last COVID-19 vaccination: Median (IQR), range | 11 (0), 10-11 |
| Prior SARS-CoV-2 infection, n (%) | 19 (86) |
| Number of SARS-CoV-2 infections in those with previous infection[1]: Median (IQR), range | 1 (1), 1-3 |
| Months since last SARS-CoV-2 infection[1]: Median (IQR), range | 50 (22), 24-68 |

[1] Missing information for one person.

### Methods

#### *Participants*

Participants were enrolled in the COVID-19 Contact (CoCo) Study (German Clinical Trials Register, DRKS00021152), an ongoing prospective observational study monitoring SARS-CoV-2 immune responses in a cohort consisting mainly of health-care professionals at Hannover Medical School (1). Participants who received 30 µg of the 2026 XFG-adapted Comirnaty vaccine in September 2026 were invited to donate blood before vaccination and 11–16 days afterwards. Of 25 vaccinated participants, three did not provide a follow-up sample within this window and were excluded from the paired analysis. The remaining 22 participants had paired samples before and at a median of 14 days after vaccination (IQR 1; range 11–16 days; table S1). None developed anti-nucleocapsid (anti-N) IgG after vaccination, and none reported SARS-CoV-2 infection between vaccination and follow-up. Forty-three percent reported underlying conditions (information missing for n=1 person), most commonly cardiovascular disease. Demographic characteristics and infection and vaccination history are summarised in table S1.

#### *Serology*

Serological analyses followed previously described protocols (2). Briefly, blood was collected into serum tubes (S-Monovette, Sarstedt, Germany), and serum was obtained by centrifugation and either analysed on the same day after storage at 4°C or archived at −80°C. SARS-CoV-2 S1-specific IgG was quantified using the anti-SARS-CoV-2 S1 IgG assay (EI 2606-9601-10G; EUROIMMUN, Lübeck, Germany) according to the manufacturer's protocol, with sample dilutions up to 1:4000. IgG against the nucleocapsid protein was assessed using the corresponding assay (EI 2606-9601-2G). Variant-resolved binding to LP.8.1 and NB.1.8.1 antigens was measured by an in-house ELISA adapted from previous work (2). Recombinant RBD and S1+S2 proteins (Sino Biological; catalogue numbers 40592-V08H160, 40589-V08H69, 40592-V08H161, and 40589-V08H71) were reconstituted at 1 µg/mL in coating buffer (BioLegend, 421701), and 100 µL was added per well to Nunc MaxiSorp plates (BioLegend, 423501) for overnight coating at 4°C. Plates were blocked with SuperBlock (Thermo Fisher, 37515) for 60 min at room temperature and washed three times with PBS containing 0·25% Tween-20. Titration curves comprised at least eight five-fold serial dilutions in SuperBlock. Starting dilutions were established empirically to achieve signal saturation and an adequate dynamic range: 1:2 for LP.8.1 and NB.1.8.1 RBD and 1:50 for the corresponding S1+S2 proteins. After 60 min at 37°C and three washes, captured IgG was detected with cross-adsorbed HRP-conjugated goat anti-human IgG (Thermo Fisher, 62-8420; 1:2000 in SuperBlock) for 60 min at room temperature. Plates were washed three times, developed for 30 min using TMB substrate (BioLegend, 421101), stopped with stop solution (BioLegend, 423001), and read at 450 nm with a 630-nm reference on a Varioskan LUX reader using SkanIt RE 6.1.1. Blank wells and an internal reference serum were included on each plate. Binding titres were expressed as ED50, defined as the reciprocal serum dilution producing half-maximal signal and derived by four-parameter logistic regression. ELISA experiments were repeated on at least three separate days using freshly coated plates and newly prepared reagents.

***Production of pseudovirus particles and pseudovirus neutralisation test (pVNT)***

Pseudovirus particles bearing SARS-CoV-2 spike proteins were generated using an established vesicular stomatitis virus (VSV) pseudotyping system (3). 293T cells were transfected by calcium phosphate precipitation with expression plasmids encoding the respective spike proteins or empty vector as a negative control. The following day, supernatants were removed, cells were washed with PBS, and VSV-G-transcomplemented VSV*ΔG (FLuc), kindly provided by Gert Zimmer, was added at a multiplicity of infection of 3. After 1 h, the inoculum was removed, cells were washed twice with PBS, and medium containing anti-VSV-G antibody (1:1000-diluted supernatant from I1 hybridoma cells; ATCC CRL-2700) was added. After 18–20 h, pseudovirus-containing supernatants were clarified by centrifugation (4000 × g for 10 min at room temperature), aliquoted, and stored at −80°C. Pseudovirus infectivity was quality-controlled on Vero cells before neutralisation testing. Spike expression plasmids represented LP.8.1 (EPI_ISL_19553000), BA.3.2.2 (EPI_ISL_19893762), NB.1.8.1 (EPI_ISL_19826033), PQ.16.1.1 (EPI_ISL_20489970), XFG (EPI_ISL_20053327), XFG.1.1 (EPI_ISL_20484922), RV.1.3 (EPI_ISL_20485417), and PJ.2.1 (EPI_ISL_20503474). Lineage and mutation information was retrieved from GISAID and CoV-Spectrum. All spike sequences were codon-optimised, cloned into pCG1, contained a C-terminal truncation of 18 amino acids, and were verified by Sanger sequencing (Microsynth SeqLab). The neutralisation assay followed a previously described protocol (4). Pseudovirus was mixed with an equal volume of serially diluted serum and incubated for 30 min before addition to confluent Vero-cell monolayers in 96-well plates. Final serum dilutions were 1:25, 1:100, 1:400, 1:1600, 1:6400, and 1:12800, with four technical replicates per sample and a no-serum control. After 16–18 h, cells were lysed with PBS containing 0·5% Tergitol for 30 min at room temperature, lysates were transferred to white 96-well plates, firefly luciferase substrate (Beetle-Juice, PJK) was added, and luminescence was recorded using a Hidex Sense microplate reader. Neutralisation was calculated from the reduction in luciferase activity relative to no-serum controls. Dose–response curves were fitted by non-linear regression to derive NT50 values. Values below the lowest tested dilution (1:25) but above 6·25 were assigned an NT50 of 25; values below 6·25 were considered negative and assigned a value of 1.

***Statistics***

Statistical analysis was conducted using GraphPad Prism 8.4 or 9.0 (GraphPad Software, USA). Outliers were included in the analysis, and missing values were excluded pairwise. Binding-antibody concentrations and ED50 values are summarised as medians with IQRs and analysed by Wilcoxon matched-pairs signed rank test. Neutralisation titres are summarised as geometric mean titres (GMTs) and Wilcoxon-Mann-Whitney-tests was used for comparison. Fold changes in the paired neutralisation panels were calculated as post-vaccination GMT divided by pre-vaccination GMT. Fold differences in the cross-variant panels were calculated relative to the LP.8.1 GMT at the same timepoint.

**Limitations of the study**

This study has several limitations. The cohort was small and extensively pre-immunised through previous vaccination and, in most participants, previous SARS-CoV-2 infection, which limits generalisability and precludes robust subgroup analyses. Serum was obtained 11–16 days after vaccination and therefore characterises an early response rather than durability (5-7); longitudinal sampling will be required to assess waning. The study did not include a contemporaneous comparator vaccine group, so quantitative differences from previous JN.1- or LP.8.1-adapted vaccine studies should be interpreted descriptively rather than as controlled comparisons. Neutralisation was assessed with a VSV-based pseudovirus assay, which is an established surrogate for authentic-virus neutralization (8), but these results require confirmation where relevant with clinical isolates. Finally, binding and neutralisation endpoints do not establish protection against infection, hospitalisation, death, or post-COVID sequelae.

**Acknowledgements**
We thank the CoCo Study participants for their support and the entire CoCo study team for help. We would like to thank J. Topal, K. Sträche, B. Heinisch, G. Baier, and A. Stölting for technical and logistical support.

**Contributions**
Study design: G.M.N.B., A.D.-J.
Data collection: C.H., M.H., M.V.S., N.C., I.N., L.G., A.-S.M.
Data curation: C.H., M.V.S, M.H.
Data analysis: M.H, C.H., M.V.S, G.M.N.B.
Data interpretation: M.H., S.P., T.W., A.D.-J., G.M.N.B.
Writing: G.M.N.B. with comments from all authors.
G.M.N.B, M.H., and C.H. have directly accessed and verified the underlying data reported in the manuscript.

**Data sharing statement**
All requests for raw and analysed data that underly the results reported in this article will be reviewed within four weeks by the CoCo Study Team, Hannover Medical School to determine whether the request is subject to confidentiality and data protection obligations. Data that can be shared will be released via a material transfer agreement.

**Ethics committee approval**
The CoCo Study (German Clinical Trial Register DRKS00021152) and the analysis conducted for this article were approved by the Internal Review Board of Hannover Medical School (institutional review board no. 8973_BO-K_2020). All study participants gave written informed consent and received no compensation.

**Competing Interests and Funding**
S.P., M.H., and I.N. declare conducting contract research (testing of vaccinee sera for neutralising activity against SARS-CoV-2) for Valneva, unrelated to this work, and S.P. served as an advisor for BioNTech, unrelated to this work. G.M.N.B. declares serving as a lecturer for Pfizer and adviser for Moderna, unrelated to this work. A.D.-J. declares serving as an advisor for Pfizer, unrelated to this work. G.M.N.B. acknowledges funding by the European Regional Development Fund ZW7-85151373 and by the German Research Foundation (Deutsche Forschungsgemeinschaft, DFG) under Germany's Excellence Strategy EXC 2155 RESIST. N.C. acknowledges funding by the Chinese Scholarship Council (202308310035). T.W. acknowledges funding by the DFG under Germany's Excellence Strategy EXC 2155 RESIST (project 390874280). S.P. acknowledges funding by the EU project UNDINE (grant agreement number 101057100), the COVID-19-Research Network Lower Saxony (COFONI) through funding from the Ministry of Science and Culture of Lower Saxony in Germany (14-76403-184, projects 7FF22, 6FF22, 10FF22), and the German Center for Infection Research (TTU 01.829). The funding sources had no role in the design and execution of the study, the writing of the manuscript and the decision to submit the manuscript for publication. The authors did not receive payment by a pharmaceutical company or other agency to write the publication. The authors were not precluded from accessing data in the study, and they accept responsibility to submit for publication.

**Suppl. References**

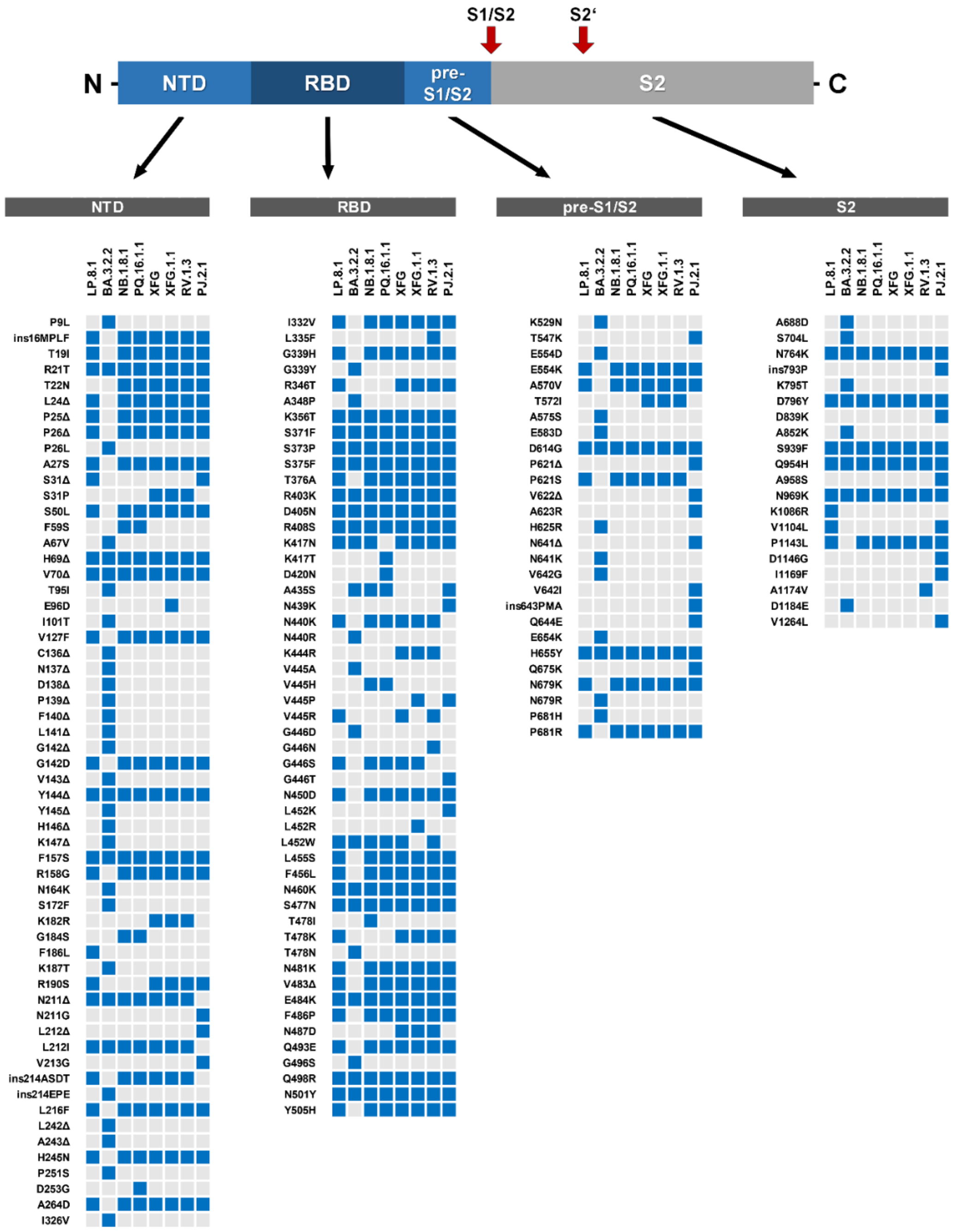

**Fig. S1 | Overview of SARS-CoV-2 lineage-specific spike protein mutations.**

Top: schematic illustration of the SARS-CoV-2 spike protein showing functional domains and cleavage sites (S1/S2 and S2′). NTD=N-terminal domain. RBD=receptor-binding domain. pre-S1/S2=region between the RBD and the S1/S2 cleavage site. S2=S2 subunit. Bottom: blue boxes indicate spike-protein mutations in LP.8.1, BA.3.2.2, NB.1.8.1, PQ.16.1.1, XFG, XFG.1.1, RV.1.3, and PJ.2.1 relative to Wuhan-Hu-1. Lineage information was retrieved from GISAID and CoV-Spectrum.

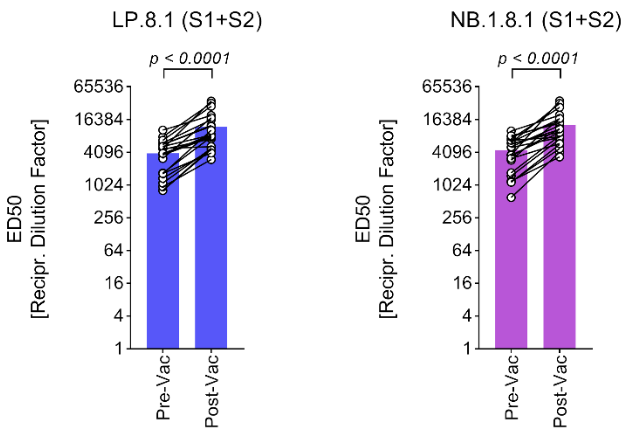


**Fig. S2 | Humoral immune responses after XFG-adapted mRNA vaccination.**

S1+S2-specific IgG-binding ED50 titres against LP.8.1 and NB.1.8.1 before and after vaccination (n=22).

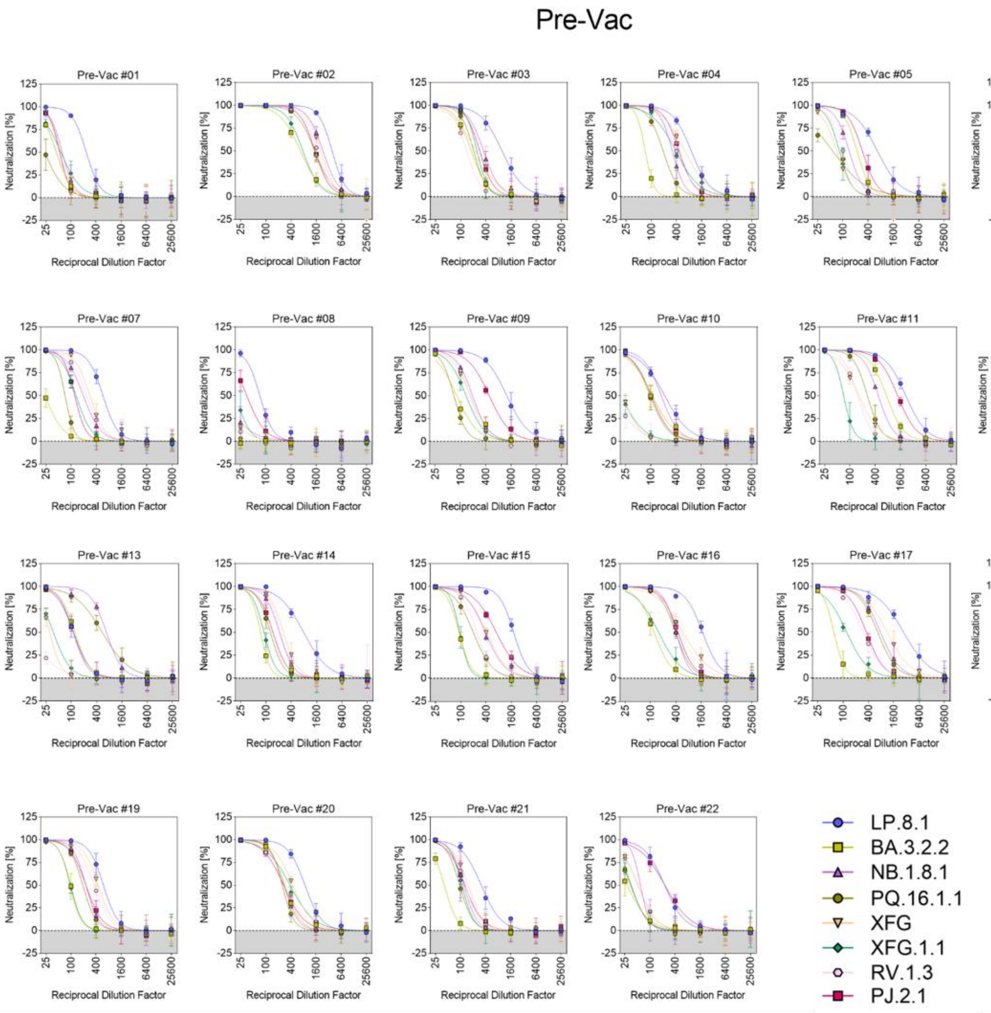


**Fig. S3 | Individual neutralisation data for pre-vaccination sera.**

Serum-specific neutralisation data for pseudovirus particles bearing the SARS-CoV-2 spike proteins under study (n=22).

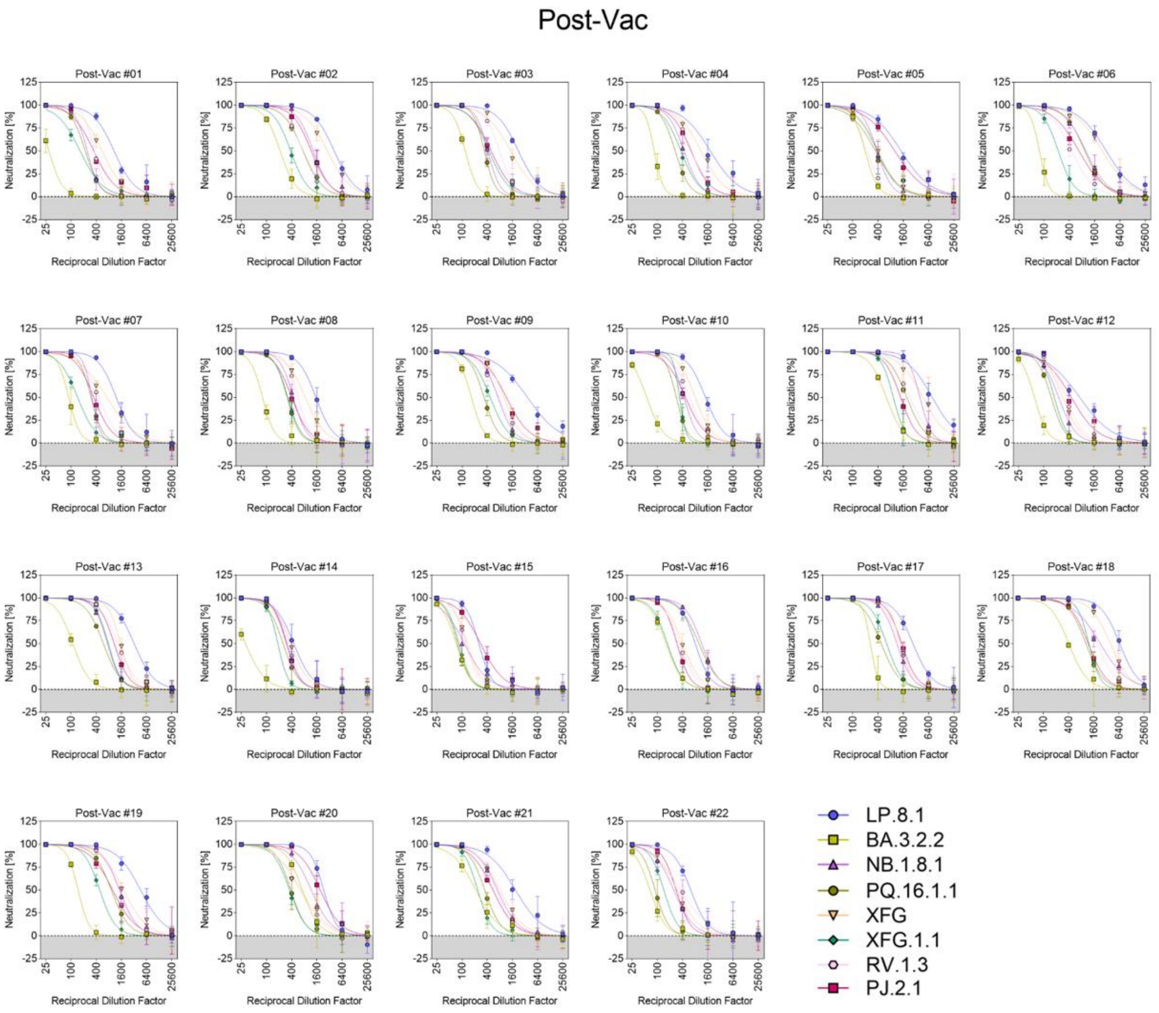


**Fig. S4 | Individual neutralisation data for post-vaccination sera.**

Serum-specific neutralisation data for pseudovirus particles bearing the SARS-CoV-2 spike proteins under study (n=22).
